# Heart Rate Assessment in a Pediatric ICU with Non-Contact Infrared Thermography and Machine Learning

**DOI:** 10.1101/2022.11.18.22282443

**Authors:** Amandeep Kaur, Samyak Prajapati, Pradeep Singh, Aditya Nagori, Rakesh Lodha, Tavpritesh Sethi

## Abstract

Heart rate is one of the vital signs for monitoring health. Non-invasive, non-contact assessment of heart rate can lead to safe and potentially telemedicine based monitoring. Thermal videos as a modality for capturing heart rate has been underexplored. Regions with large vessels such as the face can capture the pulsatile change in temperature associated with the blood flow. The use of a machine learning-based approach to capture heart rate from continuous thermal videos is currently lacking. Our present clinical investigation comprises the continuous monitoring of heart rate from a smaller number of samples by using a combination of an efficient deep-learning-based segmentation followed by domain-knowledge-based feature calculation for estimating heart rate from 124 thermal imaging videos comprising 3,628,087 frames of 65 patients, admitted to the pediatric intensive care unit at AIIMS, New Delhi. We hypothesized that periodic fluctuations of thermal intensity over the face can capture heart rate. Frequency domain features for thermal time series were extracted followed by supervised learning using a battery of models. A random forest model yielded the best results with a root mean squared error of 24.54 and mean absolute percentage error of 16.129. Clinical profiling of the model showed a wide range of clinical conditions in the admitted children with acceptable model performance. Affordable and commercially available thermal cameras establish the feasibility and cost viability of exploring deployments for patient heart rate estimation in non-invasive and non-contact environments.

## 1. Introduction

There exists multiple causes of heart rate abnormalities in infants, like fever, sepsis, anemia, shock, and pneumonia. A multitude of other pathophysiological conditions associated with heart rates in newborns as well as small children ranges from arrhythmia (irregular heart rate), and congenital heart diseases, to myocarditis and cardiomyopathy [1][2]. Low heart rates are often preterminal, and thus, recording patients’ vital signs is an essential aspect of the healthcare system. Traditional procedures, such as Electro-Cardio-Graphy (ECG) electrodes, Piezo Pulse Transducer for heart rate measurement, and Piezo Respiration Transducer for respiratory rate measurement, use uncomfortable wired measurement sensors that inflict severe limits on the patients [3]. This is true, especially for infants with fragile skin and the risk of contracting an infection through medical equipment. Doppler ultrasonography, photoplethysmography (PPG), and laser doppler sensing, on the other hand, are more advanced methods for measuring these vital signs. However, because participants in these procedures are introduced to specific active emissions, it is questionable whether prolonged use would be safe. Another disadvantage of using PPG for pulse oximetry is the likelihood of measurement error or outright failure when the patient has cold peripheries or a circulation disorder. Thus, a non-invasive, contactless method that is able to monitor vitals like heart rate is highly recommendable. These technologies are gaining momentum for their rise in application in many complex diseases such as breast cancer detection, hemodynamic shock, and skin disorders [4,5].

Computing in the modern age has revolutionized medicine with emerging technologies such as machine learning and artificial intelligence. The rapid advancements in deep learning techniques for image processing have opened a new direction for researchers to implement these techniques for contactless physiological signal extraction pipelines [6]. Contactless estimating of the pulse or respiration rate is possible by measuring variations in color and volume in superficial blood vessels during a circulatory cycle. The cause is the minuscule changes in hemoglobin concentration leading to alterations in the light absorbed by blood vessels. Despite the fact that contactless vital sign monitoring has been studied in the past, research on newborns in the Intensive Care Unit (ICU) is still restricted. Our present clinical investigation involves the constant monitoring and analysis of patients in the pediatric ICU at AIIMS, New Delhi.

Remote HR detection using thermal imaging is performed by analyzing skin temperature change, as mentioned by Garbey et al. [7]. The heat exchange by convection and conduction between vessels and surrounding tissue alters tissue temperature as a result of pulsatile blood flow. While skin color changes using optical cameras have been used to estimate heart rate, the use of thermal imaging that relies upon the accompanying change of temperature is not explored much. Sun et al. [8] proposed the first remote heart rate monitoring technique based on thermal imaging. This method computes the frequency of pulses caused by temperature variations on the vessel modulated by pulsatile blood flow. When subjected to variations in ambient temperature, one notable downside of this technique is that it may produce erroneous predictions [9].

The works of Yang et al. [10] have explored a non-contact method for heart rate and respiratory rate detection using a mid-wave thermal camera. They proposed a novel algorithm that utilizes concepts of contour segmentation to identify superficial blood vessels and performs a pixel-wise FFT for harmonic analysis. The pixels were then clustered based on their frequency components in the bands of interest, which were then processed by a median filter to create the prediction. The authors were able to attain an average RMSE of 2.83 across 20 adult patients using this approach.

Gault et al. [11] extracted the heart rate and arterial pulse waveforms from IR videos from patients under multiple physiological states. The facial region was split into 6 radial segments, centered on the nose and an axis that overlapped the eyes. Vascular maps were then created of the superficial forehead region’s arteries and supraorbital arteries of the forehead region. An algorithm was then used to identify the segmented vessels, and a specific vessel segmented was monitored to provide a 1D time-domain signal. Continuous Wavelet Transform (CWT) and Inverse CWT were used to filter out noise with a Mexican Hat wavelet, followed by additional processing to develop a wavelet-based solution. With these data, the wavelet-based approach saved computational complexity and performed better than an FFT-based approach.

This work focuses on utilizing a technique that is reliant on a purely non-invasive and non-contact procedure for extracting vitals in a critical care environment. It is important to highlight that in research environments, it is viable to use expensive equipment to prove the viability of a technique; however, in cases of actual deployment, cost-effectiveness is a factor that limits the use of such technologies. To solve this issue, this work makes use of affordable thermal cameras that are available to any clinical entity/establishment over the counter. Images from such thermal cameras were used to generate a time series of the pixel intensity measured over the region of interest (head region). This time series data was then used to generate a spectrogram in the frequency domain, and its features were extracted to train a random forest regressor to estimate its corresponding heart rate.

## 2. Materials and Methods

### 2.1 Dataset

The dataset used for training and evaluating the proposed non-invasive vital prediction pipeline is described in this section. The data collection process is described in subsection titled “Data Collection Methodology”; it discusses the various approaches used for data collection, describes the ethical approval and patient consent. Subsection titled “Data Selection and Preprocessing” elaborates on the exclusion criterions used to eliminate unsuitable videos from this study and the concluding subsection, “Data Splitting”, provides information on the unbiased methodology used for splitting data into train and test. The section titled “Methodology” delineates the methodology of this study.

#### 2.1.1 Data Collection Methodology

##### 2.1.1.1 SafeICU Framework

The data was gathered from AIIMS’s Pediatric ICU (All India Institute of Medical Sciences, New Delhi -a tertiary care hospital and the top-rated medical research university in India). The AIIMS Pediatric ICU comprises eight beds, including neonatal beds. Our servers recorded physiological vital data such as heart rate (HR), pulse rate (PR), respiratory rate (RR), blood oxygen saturation (SPO2), and other indicators in real-time. In addition, we obtained laboratory results, doctor and nurse-in-charge notes, and thermal image data. There were no constraints on the weight, gender, ethnic origin, or skin tone of the patients who were analyzed. Our current study includes 124 recording sessions from 65 UHID patients. The details of the data procurement procedure are described in the Supplementary methods S1 Section in Vanshika et al. [5] supplementary files.

##### 2.1.1.2 Ethical Approval and Patient Consent

As the data capture thermal images (infrared radiations), the identity of the infant is not revealed. In addition to this, there was no connection or variation in usual patient treatment as a result of the study. Hence a waiver of consent was sought and granted by the Institute Ethics Committee (Ref. No. IEC/NP-211/08.05.2015, AA-2/09.02.2017).

##### 2.1.1.3 Thermal Imaging Setup

The thermal camera is placed at an appropriate distance from the patient in order to avoid any direct contact or disturbance during routine check-ups as stated in Vanshika et al. [5]. The camera orientation is done in a way that captures the full body of the infant in a standard color-scaling. Videos of each patient are captured using the Seek Thermal® Compact camera (UW-AAA) with a resolution of 480 × 640 attached to an Android smartphone, with a frame rate of 15 FPS. With a 206 × 156 thermal sensor and a 36° field of view, Seek Thermal camera can detect temperatures from -40° F to 626° F. Seek Thermal camera’s cost is affordable which makes it easy for local hospitals to use this technology. The collection of data is done on different days and times so as to eliminate any bias that could occur due to the infant’s personal characteristics. At 15s intervals, vital data pertaining to the time-stamp of the videos were retrieved from the data warehouse.

##### 2.1.2 Data Selection and Preprocessing

The final data cohort was chosen through a series of procedures. Figure 1 depicts the exact pipeline followed. The videos were subjected to the first stage of review, which included checking the metadata and ensuring that all time-series data was available for the duration of the video. The videos that satisfied the above requirements progressed to the second phase of the examination. We needed at least two data points for our investigation, so only videos that met the aforementioned criteria were chosen. The third round of review included a visual screening of videos. The videos were discarded if one of the following conditions was met:

**Figure 1:**
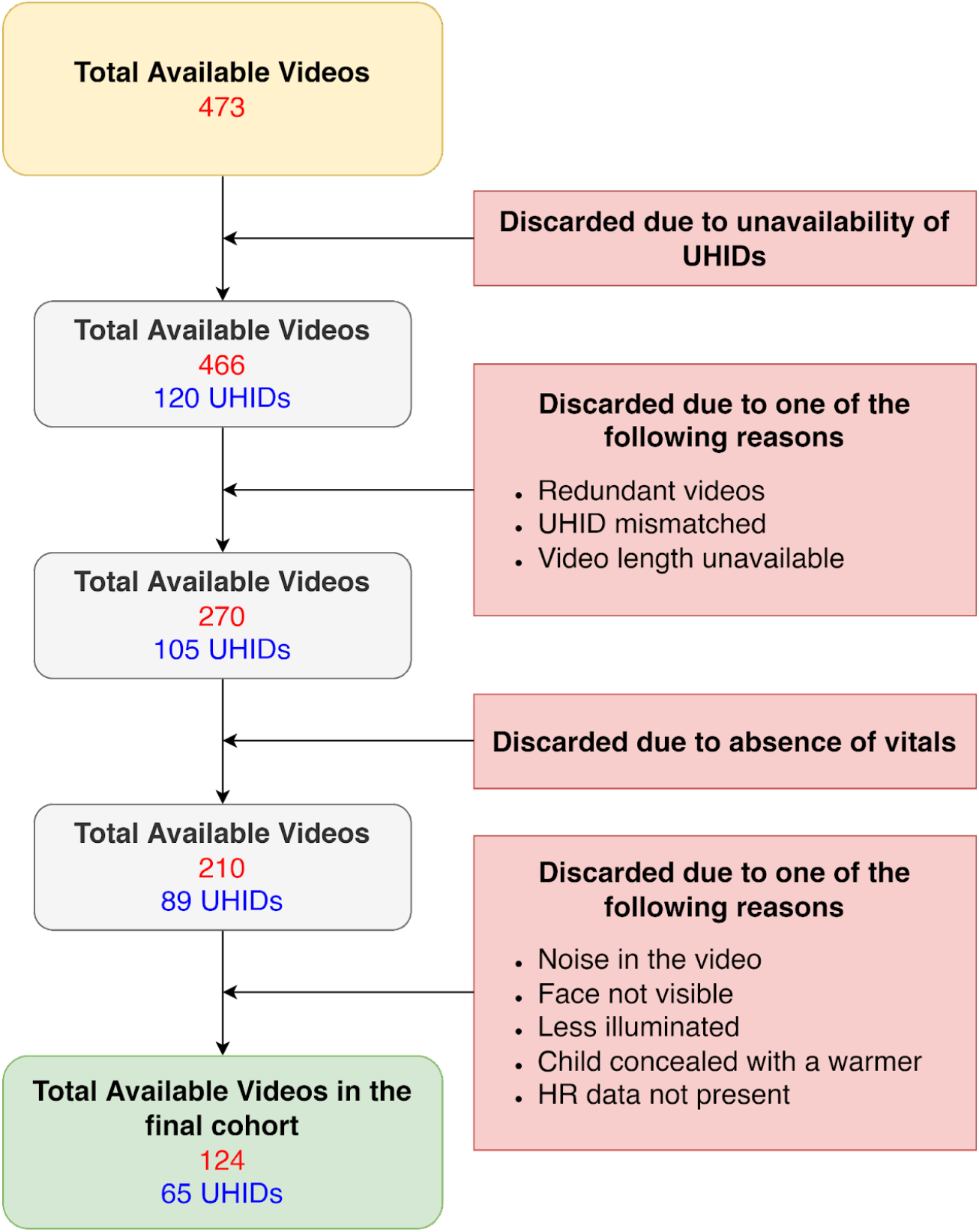
Selection criteria for data collection

- Noise in the video -If the video featured noise, the infant’s body was not clearly visible.
- Face not visible -Our analysis focuses on the facial region of the infant; hence, the videos in which the face is not visible clearly, such videos had to be discarded.
- Less illuminated -We had to eliminate certain films since the illumination was insufficient, which could lead to incorrect predictions.
- Child concealed with a warmer -This flaw was only present in a few videos.
- HR vital data missing -If NaN is present in place of HR

Finally, our dataset has 124 videos along with their time-series data for our analysis. We used the OpenCV library to decompose the videos into their respective frames at a framerate of 15 FPS.

#### 2.1.3 Data Splitting

For data splitting, we use sklearn’s StratifiedKFold. This method ensures that each fold contains a proportional distribution of data points with a given label. We make five-folds based on the ‘HR’ associated with the video. Figure 2 describes the distribution of videos within each fold.

**Figure 2:**
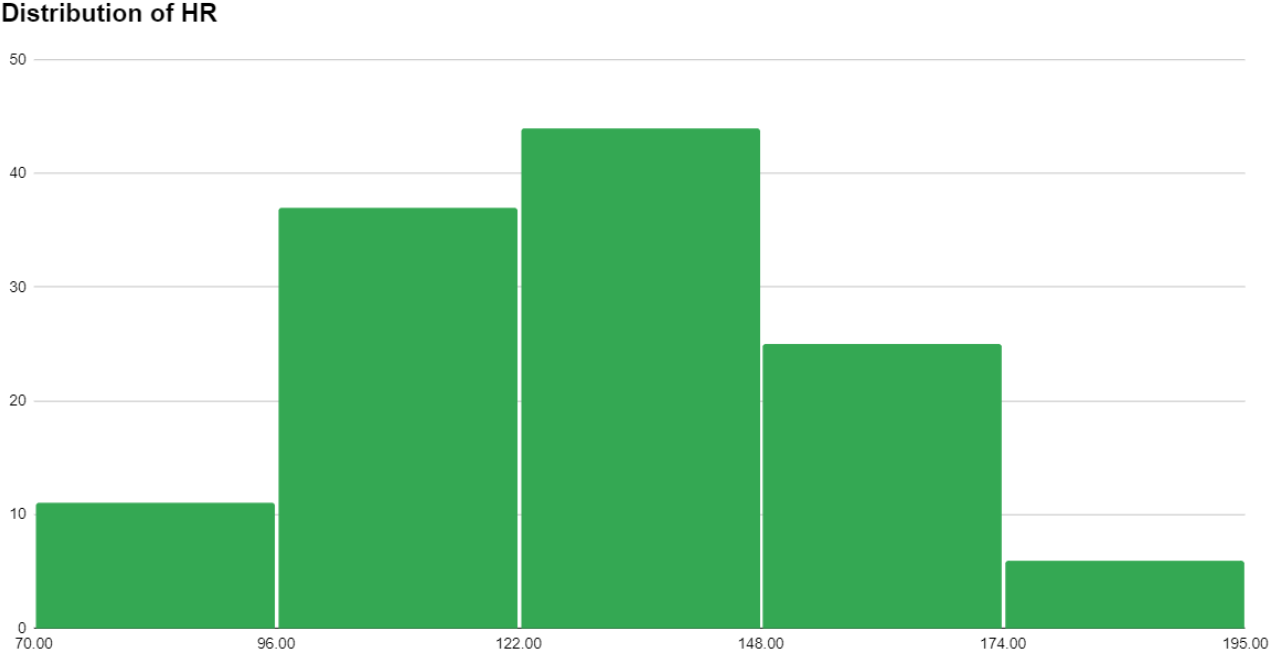
Creation of Stratified Fold on the basis of binned Heart Rate

### 2.2 Methodology

For extracting the heart rate from a single thermal video, a pipeline was proposed that enabled the extraction of spatial data about the temperature of the patient. The head region of the human body is an area that has multiple superficial blood vessels, allowing for the transmission of heat from the aforementioned blood vessels to the surface of the body [12]. A higher rate of perfusion is often followed by an increased temperature at the surface of the skin. Due to these reasons, the head region was chosen as a potential region of interest for extracting the relative temperature data.

For extracting the temperature of the head region, two object localization models were trained from scratch on detecting and localizing the head region from the previously mentioned frames that were generated by decomposing the thermal videos. This is exhibited with a pictorial representation in Figure 3.

**Figure 3:**
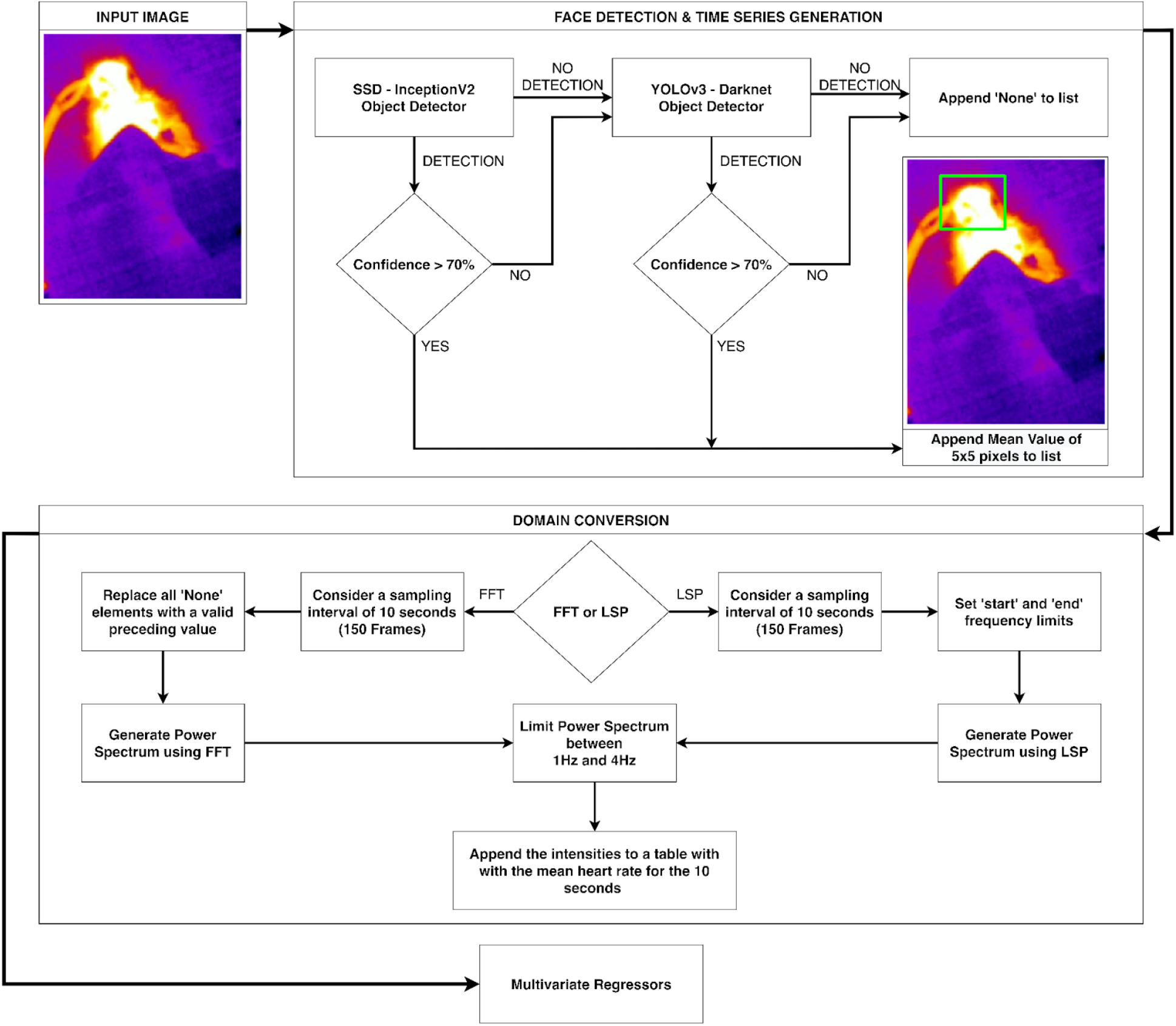
Flowchart-based description of the approach

#### 2.2.1 Face Detection and Localization

For localizing the facial region, a Single Shot MultiBox Detector (SSD) [13] based InceptionV2 was used from the Tensorflow Object Detection Library. It was trained from scratch on our SAFE-ICU data to localize the head region of neonates in thermal imagery. However, it is important to note that since these images were decomposed from videos, there exist certain frames where the Region of Interest (head region) is not visible due to movements of the body. In order to improve the detection score in such cases, the SSD-based InceptionV2 was trained with images that were augmented with randomly vertical & horizontal flips, and random image scaling as rotations. In order to further increase the robustness of the pipeline, YOLOv3 was used as a fallback algorithm, wherein in case the SSD model failed to identify the head region in an image, YOLOv3 would then be used to detect the presence of the head in said frames.

#### 2.2.2 Time Series Generation

The relative thermal data were extracted from the images by generating a one-dimensional time series from the region of interest. Certain thresholds were put into place in order to instill confidence in the object localization models.

- As discussed in previous subsection titled “Data Selection and Preprocessing”, videos were discarded if a certain video was deemed noisy, as these would lead to error-prone detections
- Non-Max Suppression was used as well with a threshold of 40% in order to eliminate multiple bounding boxes of the same region, thus improving the computational requirements.
- A confidence threshold was imposed on the models, such that a region would only be accepted to contain the head if the detection score for that region was above 70%
  - If the first object detection model is unable to detect the head region with a confidence greater than 70%, the image is then passed over to the fallback model. In case the fallback model also fails to detect the head region with appropriate confidence, it appends ‘None’ to the time series.
  - In case the 70% confidence criterion is satisfied, and the facial region is localized, its centroid is computed from the bounding boxes and an area of 5×5 pixels centered around the centroid is chosen as the ROI. An unweighted average of the 5×5 pixels was then computed for each detected frame and the value is appended to the time series.
- Since we had made use of the Lomb-Scargle Periodogram, which can work around missing values, a constraint was added from our end that guaranteed that the number of ‘None’ be less than 10% of the total values to ensure the quality of data

#### 2.2.3 Domain Conversion

The extraction of the periodic behaviors of a time series is done by using popular signal processing methods such as Fast Fourier Transform (FFT) and Lomb-Scargle Periodogram (LSP) used to convert the signal from the time domain to the frequency domain. FFT, as the name suggests, is a faster implementation of the Discrete Fourier Transform. It effectively uses a “divide-and-conquer” approach to minimize multiple redundant calculations, and due to its effectiveness at domain conversion, it is widely used in subjects requiring signal processing. However, it has certain limitations that bound it to be applicable only if certain conditions are satisfied, the major ones being,

- The sample must be evenly sampled across the entire dataset
- The highest frequency that can be analyzed depends on the Nyquist Criterion,

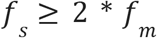

Where, f_s_ is the sampling frequency and f_m_ is the highest frequency contained in the sample, this can then be interpreted as, the highest frequency that can be analyzed is half the value of the sampling frequency.

In our work, the thermal videos were recorded at a framerate of 15 FPS, this limited our sampling frequency to 15 Hz, therefore, an upper bound was set on the highest analyzable frequency (7.5 Hz). Compounding this limitation was the fact that our object detection models sometimes failed to detect the head region in imagery, this could be due to movements of the body blurring the boundaries or simply, the head not being contained in the frame. Due to this, we opted towards using the Lomb-Scargle Periodogram for analyzing the periodicity in our signals. LSPs are most widely used in the field of astronomy where observations are subject to dips and errors due to weather, seasonal and lunar cycles, and in such cases, the sampling of data is far from even. A prominent feature of the LSP is the fact it is able to analyze signals which are sampled unevenly, and this ties in well with our use case where an averaged intensity of the ROI at any given timestamp may not be available (replaced with ‘None’ as described in the previous subsection titled “Time Series Generation”).

After generating a “master” time series of ROI intensities, a constant sampling interval had to be chosen. Intervals of 2 seconds and 10 seconds were empirically compared, and as expected, the experiment with 10 seconds as its sampling interval, performed better due to the presence of high data points. Thus, in both cases, the sample interval was chosen to be of 10 seconds (constituting of 10*15 FPS, 150 frames in each interval).

#### 2.2.4 Frequency Filtering

Since our methodology relies on a multivariate regressor to create a prediction model, increasing the number of frequencies being fed into the model increases the computational complexity of the model, and thus, increases the time taken to compute the heart rates. Thus it was imperative that we try to minimize our frequency band to speed up the computations.

The typical heart rate for a child can range from 80 to 160 beats per minute, however, medical conditions can obviously cause complications that may result in an irregular range of heartbeats. The raw vital data corresponding to each patient was analyzed, and the lowest mean heart rate was found to be 70 beats per minute, whereas, the highest captured mean heart rate was found to be 194 beats per minute. Thus, in order to capture the heart rates from the power spectrum of FFT and LSP, frequency limits were set between 1 Hz and 4 Hz, thus setting a corresponding limit of heart rate between 60 beats per minute and 240 beats per minute. This allowed us to have a certain buffer for overshoot in case the patient’s heart rate spikes.

#### 2.2.5 Execution Methodology

As discussed in the previous subsection titled “Domain Conversion”, a comparison was drawn out between the FFT and LSP-based approaches. For accomplishing this, the power spectrum intensities of each approach were fed to a multivariate Random Forest regressor in order to create a prediction model. The LSP approach performs better with reduced RMSE and MAPE values. This is attributed to the fact that FFT requires even sampling in its input data, and for satisfying this condition, each “None” value was replaced with a preceding valid value, thus duplicating the previous values to generate an even sampling. Whereas, LSP is able to work with an unevenly sampled time series, provided that the frequency limits, sampling frequency, and the “gaps” in the sample are addressed. For the final prediction of vitals signs through our pipeline, we implement five machine learning models, to narrow our search to the best performing one. We deployed Linear Regression, Decision Tree, Random Forest, SVR, and XGBoost. A series of experiments were performed for the parameter tuning. Based on the evaluation metrics, the best performing model on the given dataset was found to be Random Forest, followed by XGBoost trailing by a few decimal

### 2.3 Experimental Details

#### 2.3.1 Setup

This work is supported by a dual-socket system with 2 Intel Xeon Silver 4114 Processors and 2 NVIDIA Tesla V100 (16GB) GPUs with 128 Gigabytes of RAM. OpenCV was utilized for decomposing the videos into their individual frames at a frame rate of 15 FPS.

#### 2.3.2 Quality Assessment of Models

For the validation of our prediction model, we use RMSE and MAPE as our evaluation metrics. The Root Mean Square Error (RMSE) is defined as the standard deviation of the residuals; it shows how concentrated the data is along the line of best fit. Mathematically,

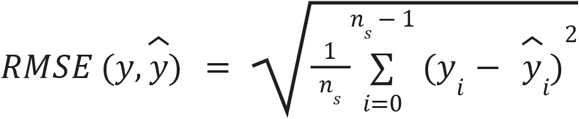

Here, *n* _*s*_ denotes the number of data points, *y* _*i*_ corresponds to the actual mean HR and 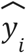 represents the predicted value.

The mean absolute percentage error (MAPE) is the average of all predicted absolute percentage errors. Mathematically,

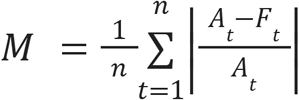

Here, *n* denotes the number of data points, *A*_*t*_ corresponds to the actual HR value, and *F*_*t*_ corresponds to the predicted vital value.

## 3. Results

Table 1 highlights the cohort description. The subjects in our research had an average age of 44 months, a heart rate of 131 bpm, and a video length of 6070 seconds. Table 2 describes the gender-specific details of patients. In our cohort of 65 patients, 47 were males and 18 were females.

**Table 1:** Cohort data characteristics and statistical significance

| Variable |  | Value |
| --- | --- | --- |
| **Age**<br>(in months) | **Median** | 26.49 |
|  | **IQR** | 66.44 |
| **HR**<br>(in bpm) | **Median** | 130.87 |
|  | **IQR** | 35.05 |
| **Video Length**<br>(in seconds) | **Median** | 63 |
|  | **IQR** | 13,872.25 |

**Table 2:**
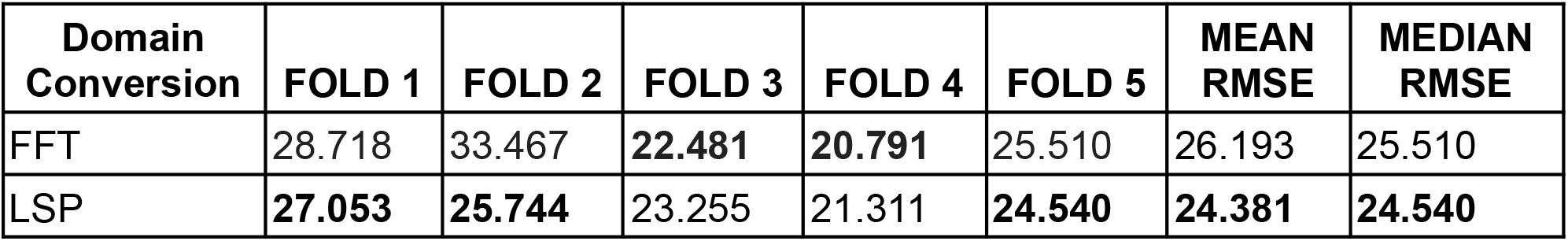
Comparison of domain conversion methods using RMSE on 5 Stratified Folds

### 3.1 Results on FFT vs Lomb Scargle comparison

Table 2 evaluates the results for both the methods using RMSE, and it is evident that LSP performs better in this scenario, with a mean RMSE of 24.381. Similarly, in Table 3, the LSP approach once again performs better with a reduced mean MAPE value of 16.873. These results are graphically depicted in Figure 4(a) for RMSE and 4(b) for MAPE.

**Table 3:** Comparison of domain conversion methods using MAPE on 5 Stratified Folds

| Domain Conversion | FOLD 1 | FOLD 2 | FOLD 3 | FOLD 4 | FOLD 5 | MEAN MAPE | MEDIAN MAPE |
| --- | --- | --- | --- | --- | --- | --- | --- |
| FFT | 19.224 | 26.628 | **13.725** | **11.205** | 17.423 | 17.641 | 17.423 |
| LSP | **19.053** | **20.599** | 16.129 | 13.699 | **14.887** | **16.873** | **16.129** |

**Figure 4:**
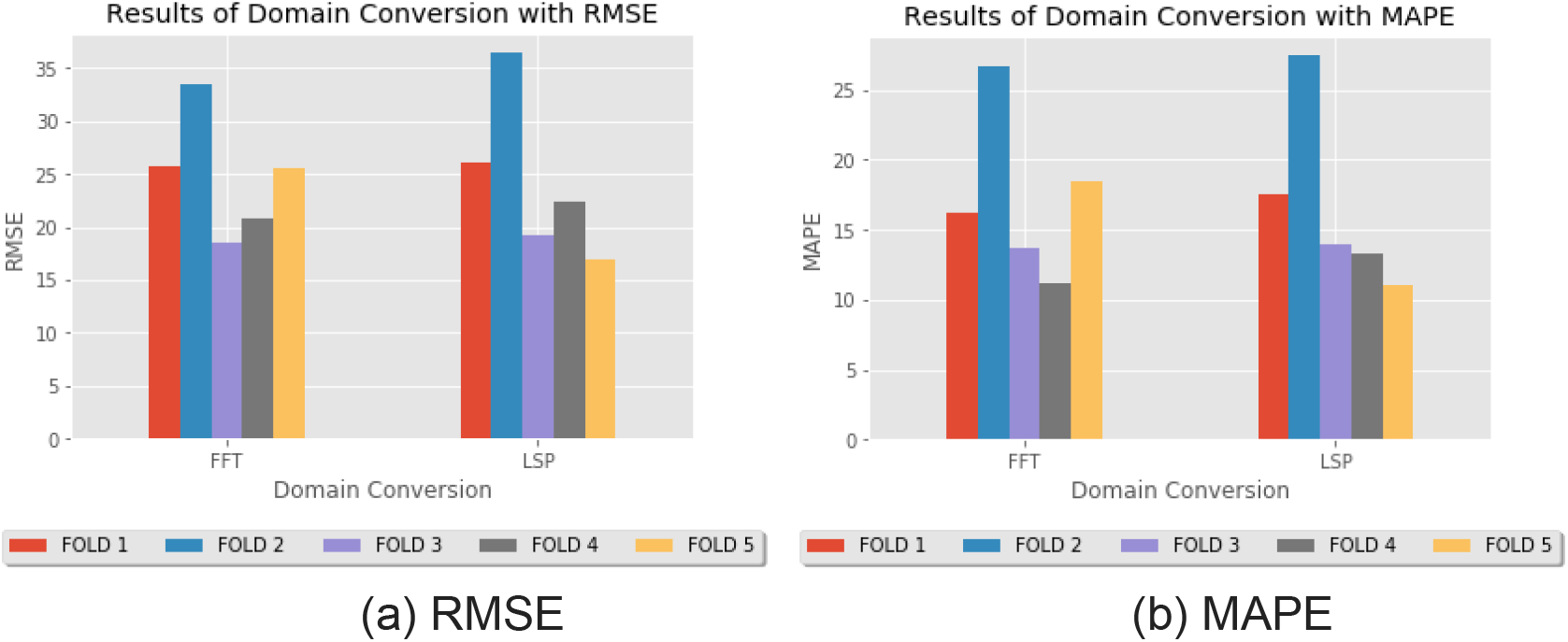
Graphical illustration of results for domain conversion methods

### 3.2 Results on different regressors on data

Table 4 displays the comparison of machine learning models using RMSE on 5 Stratified Folds. The least mean RMSE and median RMSE are obtained by the Random Forest model. Table 5 displays the comparison of machine learning models using MAPE on 5 Stratified Folds. The least mean MAPE is achieved by the Random Forest model. These results are graphically represented in Figure 5(a) for RMSE and 4(b) for MAPE.

**Table 4:** Comparison of machine learning models using RMSE on 5 Stratified Folds

| Model | FOLD 1 | FOLD 2 | FOLD 3 | FOLD 4 | FOLD 5 | MEAN RMSE | MEDIAN RMSE |
| --- | --- | --- | --- | --- | --- | --- | --- |
| **Random Forest** | **27.053** | 25.744 | 23.255 | 21.311 | **24.54** | **24.38** | **24.54** |
| **Linear** | 31.09 | **24.164** | **22.738** | 47.973 | 26.275 | 30.448 | 26.275 |
| **Decision Tree** | 36.952 | 37.902 | 35.556 | 35.12 | 36.497 | 36.405 | 36.497 |
| **SVM** | 27.967 | 25.058 | 23.752 | **20.276** | 28.772 | 25.165 | 25.058 |
| **XGBOOST** | 27.956 | 26.673 | 24.693 | 22.871 | 26.098 | 25.658 | 26.098 |

**Table 5:** Comparison of machine learning models using MAPE on 5 Stratified Folds

| Model | FOLD 1 | FOLD 2 | FOLD 3 | FOLD 4 | FOLD 5 | MEAN MAPE | MEDIAN MAPE |
| --- | --- | --- | --- | --- | --- | --- | --- |
| **Random Forest** | 19.053 | 20.599 | 16.129 | 13.699 | 14.887 | 16.873 | 16.129 |
| **Linear** | 19.599 | 19.418 | 15.285 | 18.082 | 16.337 | 17.744 | 18.082 |
| **Decision Tree** | 24.691 | 26.635 | 24.063 | 22.786 | 20.972 | 23.829 | 24.063 |
| **SVM** | 19.431 | 20.613 | 16.52 | 13.614 | 18.189 | 17.673 | 18.189 |
| **XGBOOST** | 19.591 | 20.379 | 17.071 | 14.595 | 15.618 | 17.451 | 17.071 |

**Figure 5:**
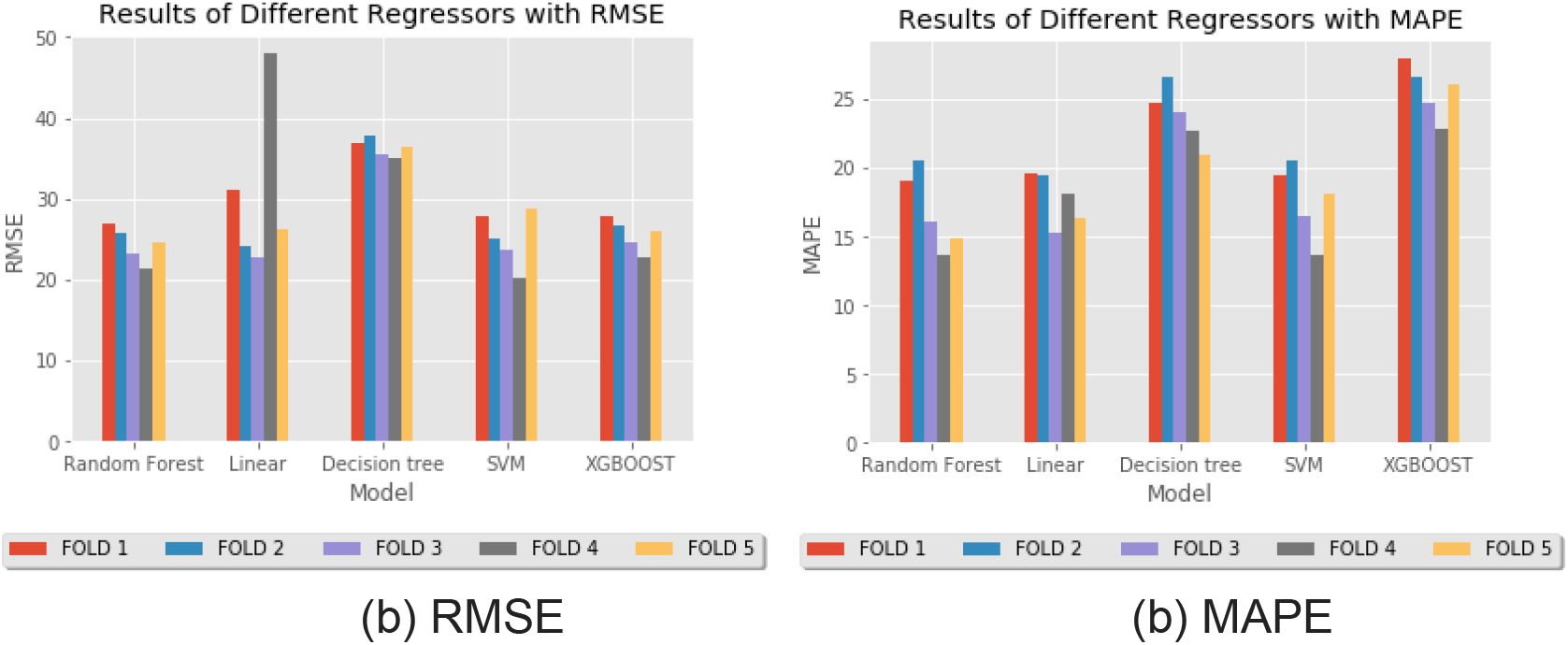
Graphical illustration of results for multivariate regressors

Clinically segmenting the cohort according to clinical conditions allowed us to examine the optimal pipeline Lomb-Scargle Periodogram with the Random Forest Regressor (LSP-RFR). In Table 6, the top 5 diseases are given along with their RMSE and MAPE values. Clinical profiling of patients with tuberculosis (13.5, 11.16) and sepsis (16.74, 10.84) had the best Mean RMSE and Mean MAPE results. This shows how broadly applicable our methodology is to critical care diseases.

**Table 6:** LSP-RFR model performance across the Top-5 diagnosis categories

| Diagnosis | Number of patients | Mean RMSE | Mean MAPE |
| --- | --- | --- | --- |
| Pneumonia | 31 | 16.71 | 11.71 |
| Shock | 24 | 19.24 | 14.4 |
| Tuberculosis | 15 | 13.5 | 11.16 |
| Acute Kidney Injury | 10 | 22.95 | 18.77 |
| Sepsis | 5 | 16.74 | 10.84 |

## 4. Discussion

In this work, we created a non-invasive computer vision-based method for extracting vitals (heart rate) in a critical care setting by analyzing 3,628,087 frames from 124 thermal videos of 65 pediatric patients from AIIMS, New Delhi, India; through this work, we were able to attain an RMSE of 24.54 and MAPE of 16.129. Head detection of neonates in thermal videos was accomplished using object detection models such as the Single Shot Detector and YOLO. A niche domain conversion method, the Lomb Scargle Periodogram was explored and it proved to give better results as opposed to the commonly used method of Fast Fourier Transform when used with the empirical outperformer (Random Forest Regressor). This is attributed to the fact that FFT requires even sampling in its data, and for satisfying this condition, each “None” value was replaced with a preceding valid value, thus duplicating the previous values to generate an even sampling. Whereas, LSP is able to work with an unevenly sampled time series, provided that the frequency limits, sampling frequency, and the “gaps” in the sample are addressed.

Methods that use Photoplethysmography (PPG) signals for extracting the vitals in works like [14,15,16,17] have a disadvantage where they require a long settling time. This could cause inconvenience to the patient under observation. Our work focuses on utilizing a technique that is reliant on a purely non-invasive and non-contact procedure for extracting vitals in a critical care environment. In our case, since the setting is contactless, there was no disturbance in the normal routine follow-ups of the patient. Conventionally, sensors required for measuring the vitals are attached to the human body using wires and require additional expensive equipment to process and amplify the signals. To combat this, the works of Yang et al. [10] use state-of-the-art thermal cameras with a temperature sensitivity of 0.025 Kelvin; cameras like these are usually bulky and expensive to procure. The feasibility of such research in actual deployments within multiple primary healthcare is deeply reduced due to the sheer cost of acquiring such thermal cameras. To solve this issue, this work relies on using thermal cameras available commercially to any clinical entity/establishment. Since each body emits radiation in the IR spectrum, using passive thermal cameras to capture, analyze and process such signals would enable us to capture variations in certain bodily functions.

It is vital that we note that our dataset was composed of patients admitted to the Pediatric ICU of AIIMS Delhi, thus, it is imperative that we recognize that each patient was diagnosed with one or more ailments (further information is present in the Supplementary section). Through our work, we have empirically concluded that the combination of the Lomb-Scargle Periodogram with a Random Forest Regressor performs the best at estimating the heart rate of a patient; we have then applied this model to cohorts of selective ailments in order to analyze the performance of heart rate estimation in that selection. Table 6 illustrates the top 5 ailments and the model performance on those ailments, and we can that the performance of the model improves when testing with a selective dataset. Further work is required in order to be certain if a model’s performance would be elevated when estimating the heart rate of a patient with a specific ailment.

It is critical to emphasize that while our dataset consisted of children hospitalized to the Pediatric ICU at AIIMS Delhi, each patient had been given a diagnosis for one or more conditions (further information is present in the Supplementary section).We have applied this model to cohorts of selected diseases in order to analyze the performance of heart rate estimation in that selection. Through our work, we have empirically determined that the Lomb-Scargle Periodogram combined with a Random Forest Regressor performs the best at estimating the heart rate of a patient. The top 5 diseases are shown in Table 6, along with how well the model performed for each disease. Clinical profiling of patients with sepsis (16.74, 10.84) and tuberculosis (13.5, 11.16) showed the best Mean RMSE and Mean MAPE outcomes. This demonstrates how generally relevant our system is to disorders requiring critical care. To be confident if a model’s performance would be improved when calculating a patient’s heart rate, more research is necessary.

As we discussed the positive parts of our experiment earlier, it is critical to discuss the limits of our research. We employ the Seek Thermal® Compact camera, which is both cost-effective and scalable, but it may fall short of providing more precise and superior high-dimensionality-based results when compared to other cameras such as the FLIR ONE. We could improve the quality of our datasets by using cameras that support higher resolution, and frame rate, and possess a higher dynamic range. This work makes use of the thermal videos from AIIMS-Delhi’s SAFE-ICU dataset, where the entire body of the child was captured at 15 FPS using a Seek Thermal Camera. This work can be refined by improving the data collection process, wherein an updated dataset can be created, which focuses on the required ROI for this study (the facial region). This would allow for a better pickup of the emissive thermal radiation generated by the superficial arteries. In the future, we want to use advanced machine learning models and deep learning models to improve performance and make it more scalable.

## 5. Conclusion

Through our study in the pediatric intensive care unit at AIIMS, New Delhi, we have demonstrated that it is possible to extract vital signals (heart rates) in a non-invasive and computationally effective way. It is critical that we note that neonates are sensitive to infections and we should ensure that patients in the Neonatal ICU (NICU) are not exposed to the unsterile environment. This study proves that research into deployments in the non-invasive and non-contact domains is explorable and economically realistic. To the best of our knowledge, this is the first study to investigate the feasibility of continuous non-invasive monitoring of pediatric thermal video data by utilizing the Lomb-Scargle Periodogram (LSP). The study investigates the monitoring of 65 infants and predicting their heart rates using a non-invasive and non-contact computer vision-based methodology. The thermal camera used in this study is affordable which could enable hyperlocal primary healthcare institutions to use this technology. Our future research would focus on using advanced computational methods for reducing errors and facilitating live clinical deployment. Prediction of cases of shock by the use of predicted heart rate is a prospective extension of this work. The present experiment on the rapid non-invasive measurement of vital signs in clinical settings has proved the immense potential of improving healthcare outcomes for people using emerging technologies that can help lower costs and increase efficiency and accuracy.

## Data Availability

All data produced in the present study are available upon reasonable request to the authors.

## 6. Acknowledgments

We gratefully acknowledge the Department of Science and Technology’s assistance with project DST/INT/ISR/P-21/2017. We’d also like to express our gratitude to Mr. Anil Sharma and Mr. Varun Prakash for their technical assistance at PICU, AIIMS, New Delhi.

## SUPPLEMENTARY SECTION

**Table S1:** Top-10 ailments in the SAFE-ICU Framework dataset cohort

| Patient Diagnosis | Incidence Count |
| --- | --- |
| Pneumonia | 31 |
| Shock | 24 |
| Tuberculosis | 15 |
| Acute Kidney Injury | 10 |
| Lower Respiratory Tract Infection | 8 |
| Ventricular Septal Defects | 6 |
| Acute Febrile Encephalopathy | 6 |
| Hypertonia | 6 |
| Gastroenteritis | 6 |
| Respiratory Failure | 5 |

## Notes

### Competing Interest Statement

The authors have declared no competing interest.

### Funding Statement

This work was supported by the Wellcome Trust/DBT India Alliance Fellowship IA/CPHE/14/1/501504 awarded to Tavpritesh Sethi.

### Author Declarations

The study was approved by the Institute Ethics Committee AIIMS, New Delhi (IEC/NP-211/08.05.2015) and involved no change in routine patient care.

